# The CREBRF diabetes-protective *rs373863828-A* allele is associated with enhanced early insulin release in men of Māori and Pacific ancestry

**DOI:** 10.1101/2021.02.27.21252567

**Authors:** Hannah J. Burden, Shannon Adams, Braydon Kulatea, Morag Wright-McNaughton, Danielle Sword, Jennifer J. Ormsbee, Conor Watene-O’Sullivan, Tony R. Merriman, Jennifer L. Knopp, J. Geoffrey Chase, Jeremy D. Krebs, Rosemary M. Hall, Lindsay D. Plank, Rinki Murphy, Peter R. Shepherd, Troy L. Merry

## Abstract

**Aim:** The minor A allele of *rs373863828* (CREBRF p.Arg457Gln) is associated with increased body mass index (BMI), but reduced risk of type 2 and gestational diabetes in Polynesian (Pacific peoples and Aotearoa New Zealand Māori) populations. This study investigates the effect of the A allele on insulin release and sensitivity in overweight/obese men without diabetes.

**Methods:** A mixed meal tolerance test was completed by 172 men (56 with the A allele) of Māori or Pacific ancestry, and 44 (24 with the A allele) had a frequently sampled intravenous glucose tolerance test and hyperinsulinemic-euglycemic clamp. Mixed linear models with covariates age, ancestry and BMI were used to analyse the association between the A allele of *rs373863828* and markers of insulin release and blood glucose regulation.

**Results:** The A allele of *rs373863828* is associated with a greater increase in plasma insulin 30 min following a meal challenge without affecting the elevation in plasma glucose or incretins GLP-1 or GIP. Consistent with this point, following an intravenous infusion of a glucose bolus, participants with an A allele had higher early (p<0.05 at 2 and 4 min) plasma insulin and C-peptide concentrations for a similar elevation in blood glucose as those homozygous for the major (G) allele. Despite increased plasma insulin, *rs373863828* genotype was not associated with a significant difference (p>0.05) in insulin sensitivity index or glucose disposal during hyperinsulinemic-euglycemic clamp.

**Conclusion/interpretation:** *rs373863828*-A allele associates with increased glucose-stimulated insulin release without affecting insulin sensitivity, suggesting that CREBRF p.Arg457Gln may increase maximal ability for β-cells to release insulin to reduce the risk of type 2 diabetes.

## Introduction

Type 2 diabetes (T2D) is a heterogeneous polygenic disease resulting from a complex interaction with environmental and genetic factors [1]. Population-specific genetic variation impacts T2D risk [2-4]. The A allele of *rs373863828*, a Arg457Gln missense variant in the *CREBRF* gene, has been associated with a 30-40% reduction in risk of T2D in people of Polynesian (Aotearoa NZ Māori and Pacific peoples) [4-6] and Oceanic (Marianas and Micronesian) [7] ancestries, as well as reduced risk of gestational diabetes in Māori and Pacific women with obesity [8]. CREBRF *rs373863828* has a minor allele frequency of 5–27% in people of Polynesian and Oceanic ancestry but is rare (<0.01%) in non-Pacific populations [4, 5, 9, 10]. The mechanism by which this genetic variant protects from diabetes is not known.

At a population level body mass index (BMI), a widely used marker for adiposity and obesity, positively correlates with risk of T2D [11] and higher BMI associates with poorer metabolic outcomes in Māori and Pacific peoples [12-14]. Paradoxically, the *rs373863828*-A allele not only associates with reduced risk of T2D, but also an increase in BMI of 1.36-1.45 kg/m^2^ (p ≤ 1 × 10^−5^) per allele [4, 5], which is the largest effect currently known of a common single variant on BMI [15]. T2D is characterised by defects in insulin secretion, insulin action, or both, resulting in chronic hyperglycemia [16]. Excess adipose tissue can reduce insulin sensitivity and impair the ability of insulin to act on peripheral tissues to dispose of glucose and supress hepatic glucose production following a meal [17].

The Gln (A) allele of CREBRF p.Arg457Gln promotes fat cell development and adiposity [4, 9], suggesting that the protective effect of the A allele on T2D may relate to preferential lipid storage in subcutaneous fat depots over metabolically more harmful visceral or ectopic fat deposition [15]. While genetically-driven metabolically-favourable fat distribution phenotypes have been reported [18, 19], there is little evidence of CREBRF Arg457Gln being a factorable adiposity allele. Indeed, the A allele associates with greater (rather than reduced) abdominal/waist circumference [4, 5], a surrogate measure for centrally located and thus metabolically unfavourable fat mass.

An alternative hypothesis for the inverse risk relationship between T2D and BMI is that higher relative muscle mass may partially account for the increased BMI, increasing the ability to dispose of glucose. Consistent with this, the A allele of *rs373863828* associates with greater lean mass in infants [20]. Both higher muscle mass and metabolically-favourable adipose tissue distribution could improve insulin sensitivity to protect from diabetes. However, whether the CREBRF Arg457Gln affects insulin sensitivity is not clear, since in people without diabetes the A allele associates with similar [7] or increased fasted insulin resistance [4] as assessed by HOMA-IR. Interestingly, however, Hanson et al. [7] reported 20% higher HOMA-β per A allele in Oceanic peoples without diabetes. HOMA-β is a marker of pancreatic β-cell function, and a decline in functional β-cell mass is a defining characteristic of T2D pathogenesis [16]. Therefore, it is possible that the CREBRF Arg457Gln variant promotes insulin sensitivity or β-cell function to maintain glucose homeostasis to slow or prevent the development of T2D.

Here we investigated the effect of the A allele of *rs373863828* on insulin sensitivity and insulin secretion in men of Māori and Pacific ancestry. We report the A allele is not associated with changes in insulin sensitivity assessed by hyperinsulinemic-euglycemic clamp. However, in response to a mixed meal challenge and intravenous glucose bolus, those with an A allele had higher plasma insulin concentration. These results suggest the CREBRF Arg457Gln variant may enhance β-cell function to protect from diabetes.

## Materials and Methods

### Participants

Healthy men of Te Moana Nui a Kiwa [Pacific Ocean; Aotearoa New Zealand Māori and/or Pacific peoples (Polynesian)] were recruited to take part in the study in either Auckland or Wellington, Aotearoa New Zealand. Informed written consent was provided, and this study was approved by the Health and Disability Ethics Committee of New Zealand (17/STH/79). Inclusion criteria were being male, aged 18-45 years, having two or more grandparents who self-report as being of Māori and/or Pacific descent [21], and being overweight or obese (BMI between 25-45kg/m^2^). Exclusion criteria included being diagnosed with a chronic illness, including cardiovascular, diabetes, and or related metabolic disease.

### Mixed Meal Tolerance Test (MMTT)

Participants were required to report to the laboratory between 0800 and 0900 hrs following a 10 hour overnight fast. They were asked to refrain from exercise, smoking and caffeine for the 24 hours prior to their visit. A cannula was placed in an antecubital vein for blood sampling, and a baseline blood sample collected prior to participants consuming a standardised mixed meal consisting of a 98 g English muffin, 13 g of butter, 23 g raspberry jam and a chocolate nutritional drink (Complan, London UK; 55 g powder in 300 ml of water) with a macronutrient break down of 71 g carbohydrate, 23 g fat and 19 g protein. Subsequent blood samples were collected at t= 30, 60, 90, 120 and 150 minutes post meal.

### Intravenous glucose tolerance test and hyperinsulinemic-euglycaemic clamp

Forty-four participants were recalled based on genotype (20 GG for *rs373863828*, and 24 AG or AA) at least two weeks following the mixed meal tolerance test to undertake a frequently sampled intravenous glucose tolerance test (FS-IVGTT) and hyperinsulinemic-euglycemic clamp [22]. All visits commenced at 0800 hrs and participants were asked to fast for 10 hrs overnight and refrained from exercise and caffeine for the previous 24 hours. Body composition was determined by dual energy x-ray absorptiometry (DXA, model iDXA, GE Healthcare, Madison, WI) and a cannula was then placed into the antecubital vein for infusion of 20% dextrose and insulin while another was placed in a wrist vein of the opposite arm for blood sampling. The blood sampling hand and wrist were placed in a heat box set at 55°C for 1 min prior to samples being taken to achieve venous blood arterialisation. Following baseline sampling a 20% dextrose bolus (0.3 g/kg, to a maximum of 30 g) was administered through the antecubital cannula within two minutes and blood samples were taken at t= 0, 2, 4, 6, 8, 10, 30 and 60 minutes. Following the 60 min blood sample, an insulin and 20% dextrose infusion were started. The priming insulin infusion dose consisted of 3 mins at 90, 75 and 60 mU/m^2^ of body surface area/min, after which insulin infusion rate was set to 45 mU/m^2^ of body surface area/min for the remainder of the clamp period. Body surface area was calculated using the Mosteller formula [23]. Blood samples were taken every 5 mins for the period of the clamp to test blood glucose (HemoCue Glucose 201+ system, Angelholm, Sweden) and the 20% dextrose infusion rate was adjusted to maintain blood glucose at 5 mmol/L. Blood samples were taken and plasma recovered at t=160, 170 and 180 mins (steady state phase) for analysis.

### Blood collection and analysis

Blood samples were collected into either EDTA or p800 (for incretins) vacutainers. EDTA tubes were centrifuged at 2000 rpm, 4°C for 10 min and p800 at 1300 rpm at 4°C for 20 mins, and the plasma was recovered and stored at -20°C until analysis. Plasma glucose, insulin, C-peptide, non-esterified fatty acids (NEFA) and triglycerides (TAG), high (HDL) and low density lipoprotein (LDL), alanine aminotransferase (ALT), C-reactive protein (CRP) and cholesterol (CHOL) were measured using Roche COBAS e 411 and c 311 analysers and commercially available COBAS elecsys assays (Roche, Mannheim, Germany). Glucagon-like peptide-1 (GLP-1) and gastric inhibitory polypeptide (GIP) were analysed using a Millipore Human Metabolic Hormone Magnetic Bead Panel (Merck, Darmstadt, Germany). A whole blood sample was taken at baseline for analysis of HbA1c (Afinion™ HbA1c, Abbott, US) and extraction of DNA. DNA was isolated using a commercially available kit (DNA extraction minikit, Qiagen, Hilden Germany). *rs373863828* genotype was determined using a custom-designed Taqman probe-set (Applied Biosystems, Foster City, CA, USA) as described previously [5, 8].

### Insulin sensitivity and secretion

Homeostatic model assessment of insulin resistance 2 (HOMA2-IR) was calculated via the Diabetes Trials Unit (Headington, Oxford, UK) online calculator described by Levy et al. [24]. Hyperinsulinemic-euglycemic clamp glucose disposal rate (GDR; glucose infusion rate (mL/hr)/ 0.3/weight (kg)) and insulin sensitivity index (ISI = GDR/((steady state insulin-fasting insulin) x steady state blood glucose) were calculated as described by Tripathy et al. [22]. Steady state blood glucose was defined as the average blood glucose at 160-180 min of the hyperinsulinemic-euglycemic clamp. Van Cauter et al. [25] kinetic parameters were used to calculate an insulin secretion curve for each subject using their measured C-peptide from the FS-IVGTT. The total area under the insulin secretion curve (AUC) from t=0 minutes to t=30 minutes was calculated as a measure of total units of secretion, the 1^st^ phase insulin secretion response from t=0 to t=4 minutes, and the 2^nd^ phase from t=5 to t=30 minutes. Disposition index was calculated as ISI from hyperinsulinemic-euglycemic clamp x average of plasma insulin at t= 2 min and 4 min of FS-IVGTT [26].

### Statistical analysis

All statistical analysis was undertaken using SPSS version 26 (IBM Corp, Chicago, IL, USA). Multivariable linear regression models were used to test for associations between *rs373863828* A allele and all parameters presented in Tables 1 and 2 with age, proportion of self-reported grandparents of Polynesian ancestry (ANC), and BMI as covariates as indicated. Statistical significance was accepted as p<0.05. Repeated measures mixed linear models were used to test associations between the A allele and all variables measured over time course experiments (MMTT and FS-IVGTT), with age, ANC and BMI as covariates. Where a statistically significant (p<0.05) main effect was seen, Sidak post-hoc analysis was undertaken, and statistical outliers were identified and removed using the Grubbs’ test.

**Table 1.** Characteristics and insulin secretion and insulin sensitivity markers in men of Polynesian ancestry with (AX) or without (GG) a A allele for *rs373863828* who underwent the mixed meal tolerance test. Association significance was assessed via multivariable linear regression with age, ancestry and body mass index as covariates.

| Variable | GG | AX | n<br>(GG/AX) | p-value | p-value<br>(BMI adj) |
| --- | --- | --- | --- | --- | --- |
| Age (y) | 28 ± 7 | 28 ± 7 | 116/56 | 0.802† |  |
| BMI (kg/m<sup>2</sup>) | 32.5 ± 4.2 | 33.1 ± 4.3 | 116/56 | 0.366† |  |
| *Fasted blood parameters* |  |  |  |  |  |
| Fasted Blood Glucose (mmol/L) | 5.6 ± 0.1 | 5.8 ± 0.2 | 115/56 | 0.402 | 0.440 |
| Fasted Plasma Insulin (μU/mL) | 13.9 ± 9.0 | 13.6 ± 10.0 | 102/55 | 0.696 | 0.537 |
| HbA1c (%) | 31.1 ± 6.8 | 30.1 ± 8.0 | 111/54 | 0.799 | 0.754 |
| HDL (mmol/L) | 1.2 ± 0.3 | 1.2 ± 0.4 | 115/56 | 0.118 | 0.115 |
| LDL (mmol/L) | 3.1 ± 1.0 | 3.2 ± 1.1 | 116/56 | 0.803 | 0.834 |
| Cholesterol (mmol/L) | 4.9 ± 1.1 | 5.0 ± 1.2 | 116/56 | 0.815 | 0.849 |
| ALT (U/L) | 25 ± 14 | 25 ± 11 | 113/55 | 0.642 | 0.620 |
| CRP (mg/L) | 1.8 ± 2.8 | 1.3 ± 1.5 | 115/56 | 0.147 | 0.138 |
| HOMA2-IR | 1.81 ± 1.14 | 1.7 ± 1.11 | 102/54 | 0.447 | 0.377 |
| HOMA2-β | 113.7 ± 65.5 | 107.7 ± 50.4 | 102/54 | 0.538 | 0.515 |

BMI, body mass index; HOMA2-IR, homeostasis model assessment 2 of insulin resistance; HOMA2-β, homeostasis model assessment 2 of β-cell function; HDL, high-density lipoproteins; LDL, low-density lipoproteins; ATL, alanine aminotransferase; CRP, c-reactive protein. Genotypes were compared using multivariate linear model with age, and ancestry as covariates. † = p-value unadjusted.

**Table 2.** Characteristics and markers of inulin release and insulin sensitivity in men of Polynesian ancestry with (AX) or without (GG) a A allele for *rs373863828* and who underwent the intravenous glucose tolerance test and hyperinsulinemic-euglycemic clamp. Association significance was assessed via multivariable linear regression with age, ancestry and body mass index as covariates.

| Variable | GG | AX | n<br>(GG/AX) | p-value | p-value<br>(BMI adj) |
| --- | --- | --- | --- | --- | --- |
| Age (y) | 29 ± 5 | 29 +/- 7 | 19/24 | 0.851† |  |
| BMI (kg/m<sup>2</sup>) | 32.3 ± 3.7 | 32.6 +/- 3.9 | 19/24 | 0.784† |  |
| Fat mass (%) | 31.4 ± 7.8 | 29.1 +/- 7.2 | 19/24 | 0.315 | 0.091 |
| Lean mass (%) | 65.9 ± 7.3 | 68.0 +/- 6.7 | 19/24 | 0.330 | 0.117 |
| *Insulin secretion modelling:* |  |  |  |  |  |
| 1<sup>st</sup> phase (AUC, mU) | 1723 ± 852 | 2464 ± 1469 | 20/23 | 0.206 | 0.193 |
| 2<sup>nd</sup> phase (AUC, mU) | 3724 ± 1355 | 3551 ± 1594 | 20/23 | 0.565 | 0.531 |
| Total (AUC, mU) | 6213 ± 1950 | 6837 ± 2746 | 20/23 | 0.745 | 0.631 |
| Disposition index | 1.37 ± 0.99 | 2.04 ± 1.94 | 20/24 | 0.130 | 0.157 |
| *Hyperinsulinemic-Euglycemic clamp* |  |  |  |  |  |
| Steady state blood glucose (mmol/L) | 5.0 ± 0.3 | 5.1 ± 0.4 | 19/24 | 0.222† |  |
| Steady state insulin (μU/mL) | 125 ± 30 | 131 ± 37 | 19/24 | 0.571† |  |
| GDR (mg/kg/min) | 7.6 ± 2.8 | 7.5 ± 3.2 | 19/24 | 0.802 | 0.903 |
| GDR (mg/kgFFM/min) | 10.8 ± 3.3 | 10.4 ± 3.6 | 19/24 | 0.905 | 0.634 |
| Insulin Sensitivity Index | 0.015 ± 0.01 | 0.014 ± 0.01 | 19/24 | 0.807 | 0.899 |

BMI, body mass index; GDR, glucose disposal rate; kgFFM, fat free mass (kg); NEFA, non-esterified fatty acids; TAG, triglycerides. Insulin secretion modelling: Van Cauter et al. [24] method applied to the FS-IVGTT to estimate insulin secretion area under the curve for 0-4 min (1<sup>st</sup> phase), 6-30 min (2<sup>nd</sup> phase) and 0-30 min (total). Genotypes were compared using multivariate linear model with age and ancestry as covariates. † = unadjusted p-value.

## Results

One hundred and seventy-two participants took part in the mixed meal tolerance test (MMTT), of which 116 had a GG (CREBRF Arg457) genotype and 56 had one or two A alleles (GA or AA; CREBRF Gln457) for *rs373863828*. Due to limited numbers of participants who were homozygous for the A allele (n=6), these participants were grouped together with those who were heterozygous (AG) and this grouping is indicated by the genotype AX. By design, GG and AX genotype groups were similar in age and BMI (Table 1). In the fasted state, those participants with the GG or AX genotypes had similar plasma glucose, insulin, HbA1c, triglyceride and cholesterol concentrations. Similarly, the presence of the A allele did not associate with significantly different circulating concentrations of the inflammatory marker CRP, the liver health marker ALT or HOMA2-IR or HOMA2-β (Table 1).

### rs373863828-A allele is associated with increased postprandial plasma insulin

There was a significant increase in blood glucose at 30 minutes following consumption of a standardised mixed meal, which returned to fasted levels by 60 minutes and was not affected by *rs373863828* genotype (Fig. 1a). Despite the similar blood glucose response to a meal, the AX genotype group had higher plasma insulin at 30 minutes following the meal (p=0.006) (Fig. 1b). Meal responses in plasma C-peptide, non-esterified fatty acids and triglycerides (Fig. 1c-e) did not differ by *rs373863828* genotype. Incretin hormones GLP-1 and GIP can potentiate β-cell insulin concentrations in response to a rise in blood glucose [27]. Given the elevated circulating insulin concentrations associated with the A allele during the mixed meal tolerance test, we investigated whether it might be due to increased concentrations of GLP-1 or GIP. While plasma GLP-1 and GIP increased (p<0.001) in response to the mixed meal challenge, it was not affected by *rs373863828* genotype (Fig. 2 a & b).

**Figure 1.**
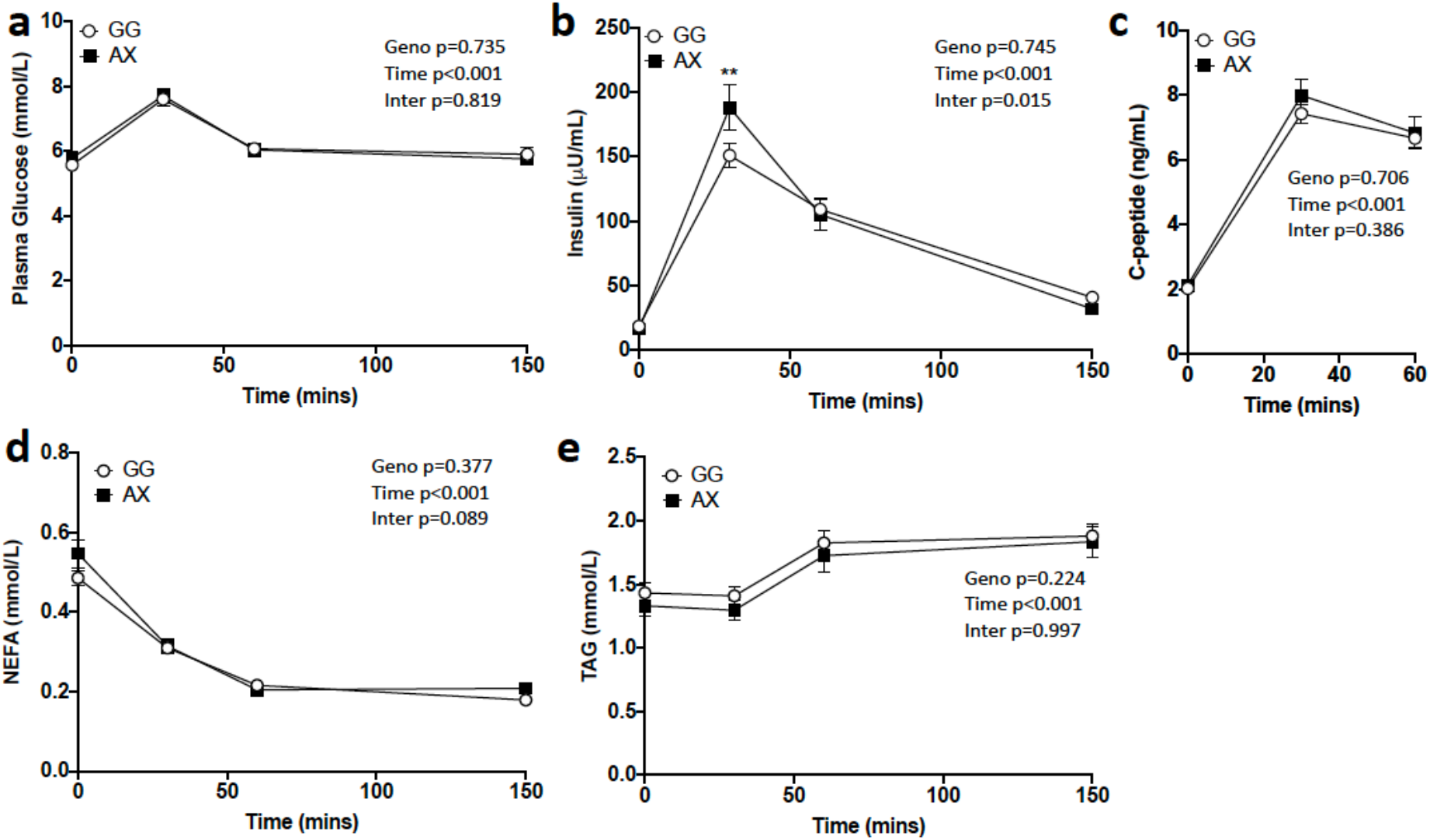
The A allele of *rs373863828* is associated with higher postprandial plasma insulin. Plasma glucose (a), insulin (b), c-peptide (c), non-esterified fatty acid (NEFA) (d) and triglyceride (e) concentration before (0 min) and following a standardised meal in men of Polynesian ancestry with (AX, n=56) or without (GG, n=113) a A allele for *rs373863828*. Data is presented as mean ± standard error of mean (SEM). Geno, Time and Inter indicate p-values for genotype, time and interaction (genotype x time) effect were assessed by mixed liner model with age, ancestry and body mass index as covariates. ** p<0.01 vs. GG at the indicated timepoint as determined by Sidak post-hoc analysis.

**Figure 2.**
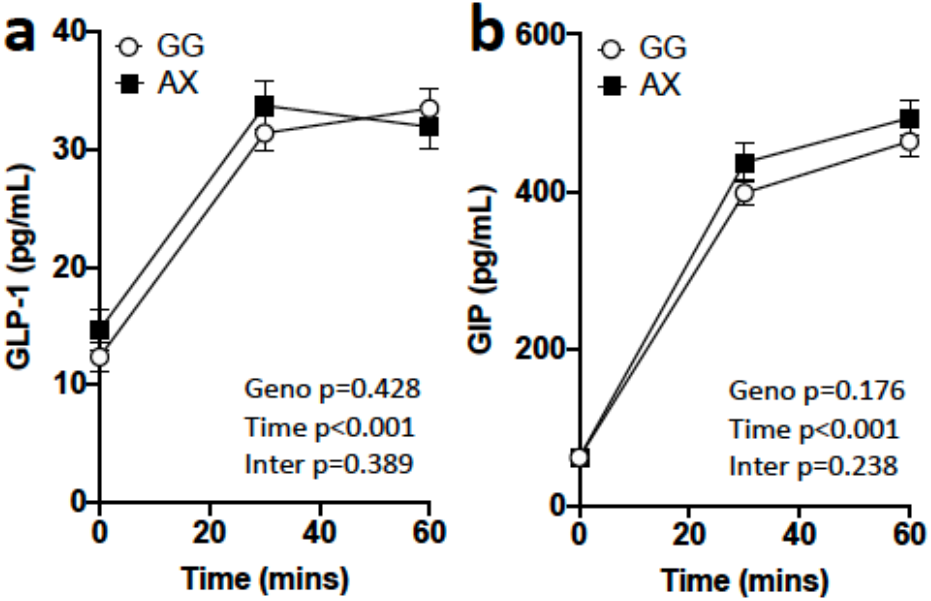
*rs373863828* genotype does not affect postprandial plasma incretins. Plasma glucagon-like peptide-1 (GLP-1) (a) and gastric inhibitor peptide (GIP) (b) concentration before (0 min) and following a standardised meal in men of Polynesian ancestry with (AX, n=53) or without (GG, n=80) an A allele for *rs373863828*. Data is presented as mean ± standard error of mean (SEM). Geno, Time and Inter indicate p-values for genotype, time and interaction (genotype x time) effect as assessed by repeated measures mixed liner model with age, ancestry and body mass index as covariates.

### rs373863828-A allele associates with increased insulin secretion in response to IV-glucose bolus

The subgroup who participated in the intravenous glucose tolerance test and hyperinsulinemic-euglycemic clamp included 20 participants with the GG and 24 with the AX genotype, who were similar in age, BMI, relative fat and lean mass (Table 2). The FS-IVGTT allows assessment of glucose-stimulated insulin secretion independent of oral absorption effects. Both genotypes had a similar peak in blood glucose concentrations of approximately 15 mmol/L during the first 2 minutes of the FS-IVGTT, with the AX genotype group having lower blood glucose at 30 min post IV-glucose bolus compared to the GG genotype group (Fig. 3a). The A allele was also associated with higher glucose-induced increase in plasma insulin and C-peptide (interaction effect p<0.05) during the FS-IVGTT, which was most evident in the 6 min following glucose bolus (Fig. 3b-c). As such, we modelled 1^st^ (0-4 min) and 2^nd^ (5-30 min) phase insulin secretion AUC using the Van Cauter et al. [25] method. Both genotype groups had similar total and 2^nd^ phase insulin secretion (p>0.50). The A allele was not significantly associated with 1^st^ phase insulin secretion (p=0.19) or disposition index (which assess the relationship between acute insulin response and insulin sensitivity [26, 28]; p=0.13), despite both variables tending to be approximately 1.4-fold higher in participants of the AX genotype (Table 2).

**Figure 3.**
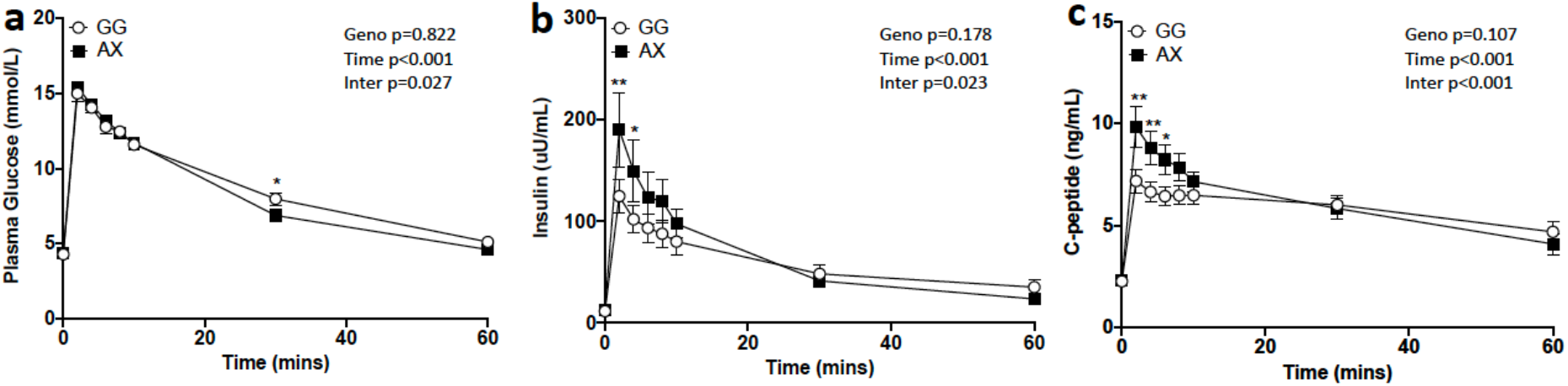
The A allele of *rs373863828* is associated with higher insulin secretion following a intravenous bolus of glucose. Blood glucose (a), and plasma insulin (b) and c-peptide before (0 min) and following an intravenous glucose bolus (0.3mg/kg) in men of Polynesian ancestry with (AX, n=24) or without (GG, n=20) a A allele for *rs373863828*. Data is presented as mean ± standard error of mean (SEM). Geno, Time and Inter indicate p-values for genotype, time and interaction (genotype x time) effect as assessed by repeated measures mixed liner model with age, ancestry and body mass index as covariates. *p<0.05, **p <0.01 vs. GG at the indicated timepoint as determined by Sidak post-hoc analysis.

### rs373863828-A allele does not associate with insulin sensitivity

Since elevated insulin concentrations for the same blood glucose concentrations in the mixed meal tolerance test can indicate insulin resistance, the hyperinsulinemic-euglycemic clamp was used to assess insulin sensitivity [22]. During the steady-state portion of the hyperinsulinemic-euglycemic clamp the two genotype groups had similar blood glucose, plasma insulin and glucose infusion rate (Table 2). Furthermore, *rs373863828* genotype did not affect whole body glucose disposal rate (GDR) or insulin sensitivity index during the clamp (Table 2).

## Discussion

The CREBRF Arg457Gln allele (*rs373863828*-A) is associated with reduced risk of T2D despite increased BMI [4, 5]. To investigate how the minor (A) allele of *rs373863828* may protect from T2D, we assessed whole body glucose homeostasis of men of Māori and Pacific ancestry using a mixed meal challenge, intravenous glucose tolerance test and hyperinsulinemic-euglycemic clamp. We found insulin sensitivity to be similar between genotype groups, however, the A allele was associated with indices of increased early insulin release in response to an IV-glucose bolus and meal challenge. This result is consistent with a report that the A allele associates with increased HOMA-β in non-diabetic Oceanic peoples [7] and suggests that CREBRF *rs373863828*-A may improve β-cell function without altering insulin sensitivity.

Glucose-stimulated insulin secretion from pancreatic β-cells is biphasic, with a rapid 1^st^ phase insulin secretion from a readily releasable pool of insulin followed by a reduced, but sustained 2^nd^ phase insulin release which continues until normoglycemia is returned [29]. Blunting of 1^st^ phase insulin response promotes postprandial hyperglycaemia and is a determinant of developing T2D [29]. While the participants in this study were normoglycemic, as assessed by HbA1c and fasting blood glucose, they were also overweight/obese (average BMI>32 mg/m^2^) [30] and had low level insulin resistance with an average HOMA2-IR index of ∼1.8 [31]. As such, it is possible this participant group had blunted early glucose-stimulated insulin secretion, and this blunting was attenuated in those with the A allele for *rs373863828*. This outcome would be consistent with the A allele protecting from T2D since preserving early phase insulin secretion is thought to be a promising T2D prevention strategy [29, 32], and restoration of early insulin release in response to weight loss is a predictor of T2D remission [33].

Plasma insulin concentrations were increased in the AX genotype group during the mixed meal tolerance test independent of changes in plasma GLP-1 and GIP, suggesting that the A allele may affect insulin release independent of gut-derived insulinotropic hormones. This outcome was supported by intravenously-infused glucose (bypassing absorption induced-gut hormone release) resulting in a greater elevation in plasma C-peptide and insulin in those with an A allele at *rs373863828*. Consistent with secretion, rather than clearance, elevating plasma insulin the AX group also had a more pronounced elevation in plasma C-peptide. C-peptide is produced when proinsulin is cleaved to insulin and is not as readily cleared from the circulation as insulin, making it a more consistent marker of secretion [34]. It is not entirely clear why insulin but not C-peptide was elevated at 30 min following a meal in the AX group. This may be the result of not capturing insulin secretion during the first 10 min following the meal, which where the genotype effect on insulin and C-peptide was most apparent during IV-GTT. Alternatively it may potentially reflect lower hepatic insulin clearance contributing to the elevated circulation insulin in the AX group following a meal [35]. However, insulin levels were similar between genotypes during the hyperinsulinemic-euglycemic clamp suggesting that any effect of the *rs373863828*-A allele on liver insulin clearance rate would most likely be subtle and/or potentially relate to a mechanism associated dependent on oral, as opposed to intravenous, glucose load [36].

Modelling of the insulin secretion profile did not detect significant differences in genotype, despite 1^st^ phase insulin secretion appearing to be 1.4-fold greater in participants with the A allele. This may be the results of a ceiling effect since we would not expect overt β-cell in this normoglycemia cohort, and therefore it is important to next assess whether *rs373863828* affects 1^st^ phase insulin secretion under conditions where there is normally a substantial suppression in early insulin release such as in pre-diabetes or early stages of T2D [29]. This may also explain why a significant genotype difference in disposition index (insulin sensitivity multiplied by the amount of insulin secreted) was not evident. The hyperbolic relationship between insulin sensitivity and insulin secretion defines the disposition index, where changes in insulin sensitivity are compensated for by inverse changes in insulin secretion and a low disposition index associates with increased risk of T2D [37-39]. However, our cohort did not fit this hyperbolic model largely owning to a limited range of insulin sensitivity and insulin secretion profiles making it difficult determine whether *rs373863828* genotype influences disposition index. It is also possible that the increased insulin levels associated with the A allele is a compensatory response to reduced peripheral tissue insulin sensitivity [16]. However, it seems unlikely given the hyperinsulinemic-euglycemic clamp showed the *rs373863828* genotype does not affect insulin-induced glucose disposal and HOMA2-IR was similar between genotypes. Furthermore, the increased insulin response in the AX genotype group associated with a greater decline in blood glucose during IV-GTT, providing further evidence that the A allele of *rs373863828* is not associating with insulin resistance.

The glucose bolus given in the IV-glucose tolerance test resulted in a >3-fold increase in blood glucose above fasting, which would likely result in near maximal glucose-stimulated insulin secretion. This may suggest that the A allele is associated with improved maximal ability for β-cells to release insulin, rather than responding to a greater glucose stimulus for release. Increased β-cell insulin secretion could be the result of improved secretory capacity or elevated β-cell mass [40]. While the effect of *rs373863828* genotype on either β-cell development, mass or secretory capacity has not been investigated, β-cell replication during infancy is a major indicator of β-cell mass in adults [41].

Recently, the A allele of *rs373863828* has been shown to associate with increased growth of lean and bone mass in the first 4-months of life in Samoan infants [20]. Whether β-cell mass follows this same trajectory has yet to be determined, but it is interesting to speculate that the A allele promotes greater β-cell mass by adulthood, and thus a greater capacity to produce insulin to control blood glucose concentration when faced with environmental factors that would otherwise promote β-cell loss and T2D. Alternatively, the A allele may promote β-cell resilience to metabolic stress-induced impairment in the internal mechanisms governing insulin secretion, dedifferentiation or death, all of which have been proposed to play a causal role in the transition from prediabetes to overt-T2D [42, 43].

CREBRF (also known as luman regulatory factor) encodes CREB3 regulatory factor protein that represses the activity of CREB3, a transmembrane transcription factor involved in coordinating the unfolded protein response (UPR) during endoplasmic reticulum (ER) stress [44-46]. While it is not known if the CREBRF p.Gln457 allele affects ER stress in β-cells, overexpression of CREBRF p.Gln457 does attenuate starvation-induced death of 3T3-L1 adipocytes [4] and elevated ER stress is one of the mechanisms through which starvation can initiate cell apoptosis [47]. Because pancreatic β-cells are inherently susceptible to ER stress-induced apoptosis and dysfunction [48], lowering ER-stress response to metabolic overload could prolong the duration in which β-cells can optimally release insulin and delay the onset of T2D.

There are a number of limitations when interpreting findings in the current study. Firstly, only men were included. Since the CREBRF p.Arg457Gln appears to increase height to a greater extent in New Zealand men of Polynesian ancestry than women [6], it is important to next assess whether this variant influences insulin secretion in females. Secondly, to avoid age and long-term disease as confounding factors, we recruited participants <45 years old and free of long-term disease. While this strategy is important to understanding whether CREBRF p.Arg457Gln alters glucose homeostasis prior to disease development, it is possible that the impact of the A allele changes with age or with development of age-related metabolic diseases and related medications. Indeed, when body composition was assessed by DXA scan, the A allele of *rs373863828* associated with increased fat mass in old age [9], but, in contrast, also increased lean mass in infants [20]. As alluded to earlier it may be under conditions where there is increased pressure on the β-cells to produce insulin (such as in prediabetes) the effect of CREBRF p.Arg457Gln on β-cells function becomes more apparent.

Collectively, our findings provide the first evidence that the Oceanic-specific A allele of *rs373863828* is associated with increased insulin release in response to a glucose challenge in overweight/obese young males of Māori and Pacific ancestry, and this appears to be independent of insulin sensitivity. We speculate that enhanced β-cell function or mass could underpin the T2D protective effect of the A allele of *rs373863828*.

## Data Availability

Considered upon request

## Acknowledgements

He mihi nui tēnei ki ngā kai tuku taonga (we would like to thank the study participants for donating their time and tissue samples). We would like to thank Prof. Vicky Cameron and Sara Raudsepp for analysing plasma incretins, and Ngati Porou Hauora for their assistance in initiating this study. This study was funded by the Auckland Medical Research Foundation (AMRF), the Health Research Council (HRC) New Zealand and the Maurice Wilkins Center (MWC). TLM is supported by a Rutherford Discovery Fellowship.

